# RObotic WAlking for children who CAnnot WAlk (RoWaCaWa): Feasibility and family impacts and perspectives of a family-led Intervention

**DOI:** 10.64898/2025.12.19.25341914

**Authors:** Jessica L. Youngblood, Benjamin M. Norman, Christa M. Diot, Karin Eldred, Sean P. Dukelow, Hana Alazem, Anna McCormick, Jennifer D. Zwicker, Patricia E. Longmuir, Kelly A. Larkin-Kaiser, Elizabeth G. Condliffe

## Abstract

**Purpose:** To evaluate the feasibility, impacts, and perspectives of a family-led robotic walking intervention.

**Materials & Methods:** This single-arm interventional study recruited participants aged ≥4 years with pediatric-onset neuromotor disorders. Participants were lent a robotic walker and recommended to use at least 150min/week for 12-weeks. Robotic walking use, acceptability, practicality and adverse events were tracked. Family goals were measured before and after training period using Canadian Occupational Performance Measure (COPM). Quality of life was examined using EQ-5D-Y, Carer-QoL, and CP-CHILD. Quantitative data were analyzed using descriptive statistics (median (25^th^-75^th^ percentile)) and Wilcoxon signed-rank tests. Qualitative interviews captured family perspectives and were analyzed using thematic analysis.

**Results:** 15 participants aged 4-23 completed this study. Participants trained 5 (3.5-6) times for 150(82-181) minutes and took 7,544(4,640 – 9,575) steps each week. Adverse events occurred in <1% (16 minor, 1 moderate) of robotic walking sessions.

Performance (3.5 (1.9-4.5), p=<0.001) and satisfaction (3.3(3.0-5.0), p=<0.001) of goals increased. Parents described positive changes in social experiences and family interactions and difficulties with the logistics of robotic walking.

**Conclusions:** This family-led robotic walking intervention resulted in improvements in individual goals, though families did struggle with some logistics of robotic walking, such as; transport and difficulties with the device.

## Introduction

It’s estimated that approximately 1/1000 children are unable to walk due to cerebral palsy, and there are many others who are unable to walk due to other diagnoses[1], [2] This inability to walk impacts almost all aspects of their lives and increases the need for caregiver support. Further, being unable to walk independently often leads to a sedentary lifestyle, which can lead to other health sequelae impacting quality of life [3] However, there are currently very few evidence-based interventions for individuals with significant motor impairments who are unable to walk in community settings. Robotic walking is a promising new technology to increase upright mobility in this population; however, the feasibility of its implementation by families and caregivers in a home and community settings remains unknown.

Interventions delivered in a home and community setting offer advantages and can address some of the challenges associated with, delivering interventions in clinical settings. Home interventions facilitate increased rehabilitation intensity and have been associated with positive family and social impacts [4], [5]. Further, they give parents autonomy to choose when activities are convenient for them and require fewer changes to their schedule than attending appointments [5]. However, the majority of robotic walking interventions have been conducted in a clinical setting. This can create barriers to care and put pressure on the healthcare system by taking clinicians’ time and other resources [6], [7] In a traditional clinical setting, children are unable to train with the same frequency as at-home interventions due to the location of the clinic, lack of clinicians’ time, financial barriers, and long wait times [5], [6], [8]. Even within an intervention session, therapists experience limited resources and long setup times [7].

Walking with an assistive device and robotic walking specifically have been shown to have broad positive impacts. Walking with a manual walker in community settings has been shown to make children who cannot walk without an assistive device happier, improve their self-confidence and independence and promote participation [9] Individuals who have greater mobility impairments are unable to experience this. However, children demonstrated pride and pushed themselves further while robotic walking in a clinical setting with parental involvement, suggesting robotic walking may have similar benefits [8] The majority of current robotic walking literature in both children and adults focuses on physical and walking outcomes [[10], [11], [12]. Therefore, the current literature may not report many of the potential benefits of robotic walking, including changes in quality of life. Though there is limited focus on patient-oriented outcomes, studies have shown significant improvements in both performance and satisfaction using the Canadian Occupational Performance Measure (COPM) [13]. Despite this promising early evidence of psychological and physical benefits, there are limited opportunities for this population to experience walking.

A robotic walker can facilitate walking for children with severe mobility impairments and be used outside of a clinical setting, such as at home, school, outdoors and in other areas of the community [14]. Most robotic walking aids are designed for a clinical setting or use by individuals who are able to ambulate without the use of a wheelchair[9], [15]. Therefore, there is a need for an intervention examining the impacts of at-home robotic walking for children with severe mobility impairments. Robotic walking has been shown to improve walking speed and endurance, gait, gross motor function, and oxygen consumption [16], [17], [18]. Though the potential benefits of using these devices at home remain relatively unknown. Previous case and observational studies involving a robotic walker used by children in community settings support that children can increase stepping activity and experience improvements in sleep, bowel function, head control, and spasticity, all of which may decrease the need for caregiver support [19], [20], [21]. Training at home with a robotic walker may impact the family differently than training in a clinical environment, such as decreased parental burden, increased engagement of the child, fostering increased motivation compared to other rehabilitation therapies and increased opportunities to experience walking together [4], [5].

The purpose of this study was: 1) To identify if a 12-week at-home robotic walking intervention is feasible for families with a child with a mobility impairment, 2) To understand the impacts on family goals and perspectives following a 12-week at-home robotic walking intervention.

## Methods

### Study Design

A single-arm, mixed-methods interventional study was conducted to assess the feasibility and outcomes of robotic-assisted gait training using the Trexo Plus (Trexo) robotic walker over a 12-week period in participants’ home and community environments. We employed a triangulation mixed methods approach in order to understand the experience of individuals through a variety of data collection methods. Ethics approval was obtained from the University of Calgary Conjoint Health and Research Ethics Board (REB21-1166), and written informed consent was obtained from all participants’ legal guardians. The trial was registered with ClinicalTrials.gov (ID: NCT05473676).

### Device Description

Participants used the Trexo robotic walker to complete robotic walking sessions in a community setting. This system consists of bilateral robotic legs attached to a four-wheeled walker and is uniquely designed for use in community settings [12]. The device supports users up to 150 lbs with femur lengths <35 cm and calf lengths <43 cm. It is adaptable to individual user needs, allowing configuration with or without the Rifton saddle, and can accommodate both anterior-facing and posterior-facing positions (depending on the user’s trunk control. The motors are aligned with the hips and knees and facilitate ambulation through a pre-programmed gait cycle tailored to each participant’s joint range of motion [12]. Device parameters—including joint range of motion, motor force limits, and cadence—were configured via a tablet interface connected over Wi-Fi. The system requires a caregiver or trained assistant to accompany the user during operation to guide steering and monitor tablet control [12,18]. A total of six Trexo robotic walkers were made available throughout the study, including three large Trexos paired with large Rifton Pacer frames, two medium-sized Trexos with large frames, and one medium-sized unit with a medium Rifton Pacer base. Allocation was based on participant size and availability across timepoints in the study.

### Recruitment and Participants

Participants could be referred by their treating clinician or self-refer in response to study information shared through social media, clinical teams, or community members or organizations (i.e. convenience sampling). Interested families, completed a trial to confirm they met inclusion criteria and the size of Trexo needed. Inclusion criteria were: unable to walk independently due to pediatric-onset, non-progressive central nervous system disorder or injury, at least four years of age, able to fit in a Trexo (have a thigh measurement <34cm, lower leg measurement <42cm and weight less than 150lbs), and were able to comply with study procedures (assessments and training). Participants were excluded if at the time of the study they had any one of the following: a medical condition or recent surgery requiring physical activity restriction (eg. unstable arrhythmia) or lower extremity immobilization or weight-bearing restrictions (eg. fracture, unstable hip subluxation), pain or symptomatic hypotension while standing, contracture such that the Trexo robotic gait trainer does not result in forward movement, or contemporaneous involvement in a potentially confounding intervention. Initially, participants who expressed interest after trialling the device and met all inclusion criteria were enrolled. When enrollment demand exceeded device availability, participants were randomly selected from the list of eligible individuals who fit the device size available next.

### Intervention and Procedure

This study intervention involved lending families a Trexo robotic walker for 12 weeks to use in their homes and other community settings. Participants were asked to use the robotic walker for at least 150 minutes/week (30 minutes/day, 5 days/week), consistent with current rehabilitation recommendations to see changes in neuroplasticity [22]. Caregivers who ran the robotic walking sessions throughout the study received training in a three-hour Zoom call with Trexo Robotics. During this training, the caregivers learned how to put the participant in the Trexo and make the adjustments necessary for the participant’s comfort, use the tablet and run/connect the Trexo. A member of the research team had periodic (weekly to monthly, depending on participant need) check-ins to support logistics and adherence to the intervention frequency.

Participants were enrolled for a baseline period of at least 4 weeks before the robotic walking intervention to enable the collection of multiple baseline values (see Figure 2) and followed throughout a 12-week intervention and subsequent 12-week follow-up period. Interested families who were identified at least three months prior to device availability were asked if they were willing to participate in additional assessments three months before the intervention start date, to generate pilot data on the stability of the outcome measures. Figure 1 represents when each outcome measure was collected. Additional outcome measures examining the body function of our participants were collected as part of this study; those outcomes will be reported elsewhere.

**Figure 1:**
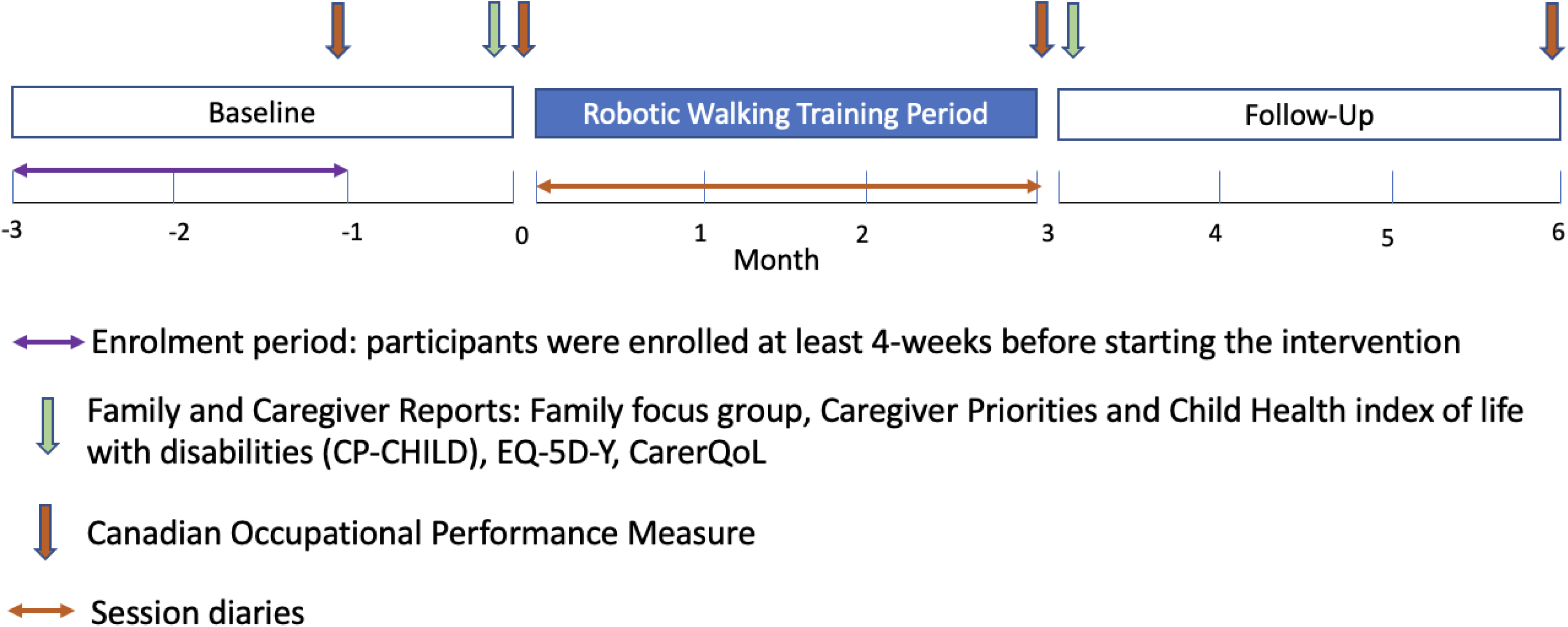
Timeline of study and when all outcomes were collected

**Figure 2:**
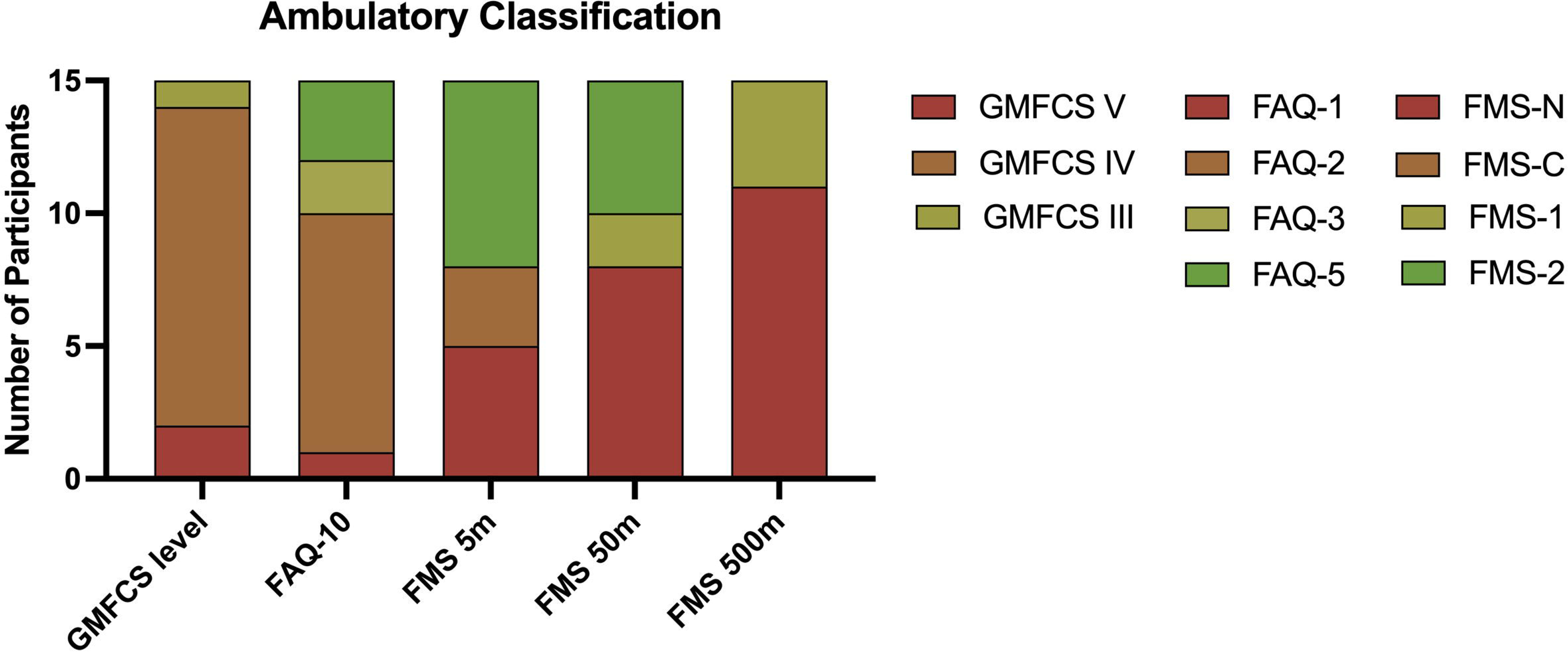
Ambulatory ability of participants. Ambulatory ability was classified through the Gross Motor Function Classification System (GMFCS), Gillette Functional Ability Questionnaire (FAQ), and the Functional Mobility Scale (FMS).

### Quantitative Methodology

#### Outcome Measures

A parent or guardian completed demographic questionnaires and outcome measures, and when possible, participants who were capable were also welcome to participate in reporting.

##### Feasibility Outcomes

Feasibility was assessed through six categories (retention, demographics, adherence, acceptability, practicality, and safety) to gain a more complete understanding of the feasibility of this intervention for families [5].

Retention was assessed by the number of participants who completed the study, as well as understanding the reasons participants were unable to complete the study.

A parent/guardian filled out a demographics questionnaire when they were first enrolled in the study. This questionnaire asked the child’s date of birth, sex, gender, underlying cause of mobility impairment, Gross Motor Function Classification System (GMFCS) Level [23], [24], first language, the Gillette Functional Assessment Questionnaire (FAQ) [25] and Functional Mobility Scale (FMS) [26]. While the GMFCS is designed and validated for use in individuals with cerebral palsy, the definition of cerebral palsy is not perfectly consistent, and it is a common communication tool used in the CP-like conditions that were represented by our participants [27]. We also asked for additional information regarding the family’s dynamics, ethnicity, and socioeconomic status to gain a better understanding of who this intervention was feasible for.

Adherence was assessed by the participants’ usage (defined as meeting training targets of 150 min/week) and the number of steps taken in a week. The number of steps taken and time used were accessed from Trexo’s database, where they upload all usage information collected from the Trexo tablet. There is a data sharing agreement in place which allowed us to access this information. We also looked at the different demographic and training parameters that may have impacted participants’ ability to meet training targets. Anything impacting training adherence was also discussed during the check-ins with the research team.

Daily diaries were filled out every day of the intervention period (before & after each training session and once on rest days). These diaries tracked outcomes related to acceptability, practicality and adverse events. Acceptability was assessed through investigating if training impacted the participant’s mood and asking about the caregiver’s experience. The parent/caregiver running the robotic walking session rated the participant’s mood on a Likert scale of 1-5 (Very Bad = 1, Bad = 2, Okay = 3, Good = 4, Very Good = 5) before and during each robotic walking session to determine if training resulted in a change in mood. The caregiver were asked to rate their experience during the training the same Likert Scale. The average scores across all training sessions was used for each participant combining all parent/caregivers working with that participant.

Practicality was assessed in multiple ways. The session diaries captured where they were robotic walking (options given: home, school, outside, recreation center, other), who ran each session (options given: parent, aide, other) and any problems that occurred during the session (options given: malfunction with the device, difficulty with set up, lack of time, child not enjoying or not interested in training, other). Finally, practicality was also examined through the semi-structured qualitative interviews with parents.

Safety information was obtained during check-ins when parents disclosed that there was an adverse event. Session diaries were also used to capture adverse events that occurred during the training period. Adverse events were then classified as any event related to robotic walking or events not related to the device, such as unrelated injuries or illnesses. Adverse events related to robotic walking were then classified as mild (slight discomfort, requires no practitioner support), moderate (need to see practitioner but not a prolonged problem), and severe (requires in-person hospitalization and results in persistent injury).

##### Family Impacts and Goals

Family goals were identified and evaluated using the Canadian Occupational Performance Measure (COPM) [28]. Clinicians guided a discussion with the family about areas of the participants’ daily life to help the family identify specific goals that were meaningful to the family and would possibly be impacted by robotic walking. Parents then rated their child’s Performance and their Satisfaction with the current performance of each goal on a scale from 1-10. These ratings were collected 4 weeks before and directly prior to intervention, directly following intervention, and 12 weeks after intervention (see figure 1). Parent goals were categorized to understand goals that were important to families. These categories were: Functional mobility (e.g. walking, crawling, transfers), personal care(e.g. dressing, bathing, feeding), socialization (e.g. communication and peer interactions), active recreation (e.g. sports and other physical activity), school and/or play (e.g. play skills, school activities, homework), quiet recreation (e.g. hobbies, watching tv, reading). (Law et al, 2004; Walsh et al, 2024)

Three questionnaires addressing the health and quality of life of the participants and their parents/guardians were filled out by parents directly before and after the intervention period. The Caregiver Priorities and Child Health Index of Life with Disabilities (CP-CHILD) questionnaire is a reliable and validated measure to understand the caregiver’s perspective of activity limitations, health status, well-being, and ease of care for their child living with cerebral palsy who are GMFCS III-V [29] The EQ-5D-Y was developed to assess health-related quality of life in children and adolescents [30] The final questionnaire filled out was the Carer-Qol, which examines caregiver quality of life and is used to assess the burden experienced by informal caregivers when taking care of individuals with a disability [31].

##### Quantitative Analysis

The average usage/week (minutes, steps, times used and cadence) was calculated for each participant. Given the diversity of our small sample, we present non-parametric descriptive statistics (median (25th – 75th percentiles)) for all outcome measures. A Friedman’s Correlation was used to test for changes in usage parameters across the study period. A Wilcoxon signed rank test of difference was used for pre to post and pre to 12-week follow-up measures to compare if there was a change in scores when comparing these timepoints. An alpha of 0.05 was used for all outcome measures.

##### Qualitative Methodology

###### Theoretical Framework

The qualitative aspect of this study was guided by a relativist ontology and constructivist epistemology. A relativist ontology assumes that reality is constructed by lived experiences and therefore each human has constructed their own reality. This ontology acknowledges that multiple realities exist [32]. A constructivist epistemology acknowledges that knowledge is gained through interaction with and understanding the experiences of others [32]. This philosophical approach allowed the researchers to build knowledge about the commonalities and differences in the experiences of families as they relate to using the Trexo device, recognizing that each family will have had different experiences and interactions all of which will be reflected in their “truth”.

###### Methodology

Qualitative description is the overarching framework used for this study. This methodology is a general qualitative approach used in mixed-methods health research to collect and interpret first-hand experiences from knowledge users. One of the main tenets of this approach is to stay close to the data to create a rich description of how knowledge users experienced an event. Like most qualitative methodologies, this approach is inductive in nature to describe participants’ experiences that address the research question [33], [34].

##### Data Collection

Semi-structured interviews were used to further understand parents’ perspectives on the feasibility of this intervention and to understand the impact this intervention had on families. Interviews were conducted with parents or guardians, and participants themselves were also invited to participate if they were both able and interested. The pre-training interview guide focused on what the family wanted to get out of robotic walking and areas of their life where robotic walking may help, as well as other thoughts and feelings they have regarding the device. The post-training interviews focused on questions regarding the feasibility of using the device in their home communities, the impacts robotic walking had on the child, and whether pre-training expectations were met. Interviews were offered via videoconference (Zoom) or in-person, and all participants chose videoconference.

### Qualitative Analysis

NVivo12 software (QRS International Pty Ltd.) was used to store and analyze data. Interviews were audio recorded and transcribed verbatim by Zoom. All transcripts were anonymized, read and compared to the audio recordings to ensure transcripts are correct. This data was analyzed from the perspective of our relativist ontology and constructivist epistemology. Thematic analysis was conducted to group and understand themes and patterns across the dataset and allowing for the comparison and contrast of family experiences. Thematic analysis was conducted in six phases: 1) Transcripts were read and re-read to gain familiarization with the dataset; 2) The transcripts were then read through to generate initial codes; 3) Similar codes were grouped to create themes across the dataset; 4) Themes were then reviewed and checked to ensure they align with the research question; 5) Names and definitions of themes were created; 6) Finally, the report was written [35], [36]. This analysis method is particularly useful for this study, as it allows researchers to compare and contrast all the parents’ perspectives and therefore gain a wide view regarding families’ perspectives on the feasibility and family impact of robotic walking.

#### Qualitative Rigour

The rigour criteria for this study were assessed against criteria that align with our relativist ontology, constructivist epistemology and reflexive thematic analysis approach. The first author kept a reflexivity journal where she reflected on my interpretations of the data and how her involvement impacted the research process. Resonance occurs when the findings are transferable, and the reader can make connections between the findings and their lived experience. Resonance was addressed through thick description, where findings are thoroughly discussed and provide contextual information so the reader can make connections to their own lived experience [37].

## Results

### Feasibility

#### Retention

There were nineteen participants enrolled in this study, and fifteen who completed the intervention. One participant withdrew before training started as they were offered a date during the intervention period for a needed orthopedic surgery. The additional three participants withdrew within the first month of the training period. One participant had a significant illness and, afterwards, the family decided it was no longer feasible for them to participate. The final two withdrew because they had planned to use the robotic walker at a school, but a policy changed, so it was no longer possible to train at school, and it was not feasible for the families to do the training at home.

#### Demographics

The participants who completed the study were aged 6.5 (5.3 −10) years, and the majority of participants (10/15) were diagnosed with cerebral palsy (Table 1). The majority of participants were unable to walk independently (FMS 2), had difficulties sitting without support and had some mobility, such as crawling or using a powered wheelchair (GMFCS IV) (Figure 2). All participants had bilateral motor impairments, with most (14/15) having quadriparesis. Of the participants who had Cerebral Palsy, all had spasticity; five had spastic-dyskinetic hypertonicity, four had primarily dystonia, and one had athetosis motor impairments. The remaining five participants had genetic conditions and experienced the following: three had hypotonia, and two had spastic-dyskinetic motor impairments. 11/15 participants were white, two were Latin American, one was Indigenous, and one was East Asian and white.

**Table 1:**
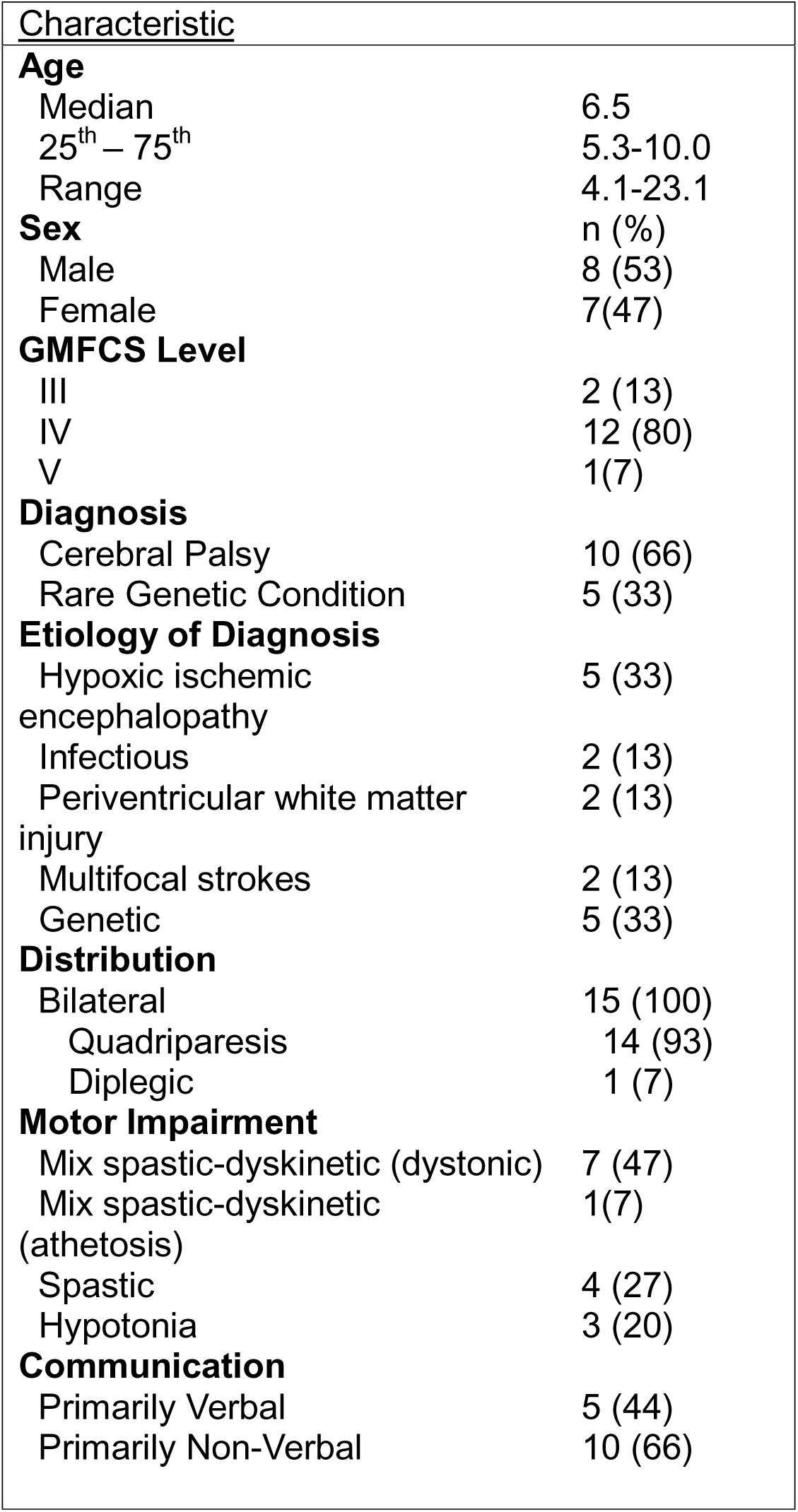
Participant Characteristics.

| Characteristic |
| --- |

|  |  |
| --- | --- |
| **Age** |  |
| Median | 6.5 |
| 25<sup>th</sup> – 75<sup>th</sup> | 5.3-10.0 |
| Range | 4.1-23.1 |
| **Sex** |  |
|  | n (%) |
| Male | 8 (53) |
| Female | 7(47) |
| **GMFCS Level** |  |
| III | 2 (13) |
| IV | 12 (80) |
| V | 1(7) |
| **Diagnosis** |  |
| Cerebral Palsy | 10 (66) |
| Rare Genetic Condition | 5 (33) |
| **Etiology of Diagnosis** |  |
| Hypoxic ischemic encephalopathy | 5 (33) |
| Infectious | 2 (13) |
| Periventricular white matter injury | 2 (13) |
| Multifocal strokes | 2 (13) |
| Genetic | 5 (33) |
| **Distribution** |  |
| Bilateral | 15 (100) |
| Quadripareisis | 14 (93) |
| Diplegic | 1 (7) |
| **Motor Impairment** |  |
| Mix spastic-dyskinetic (dystonic) | 7 (47) |
| Mix spastic-dyskinetic (athetosis) | 1(7) |
| Spastic | 4 (27) |
| Hypotonia | 3 (20) |
| **Communication** |  |
| Primarily Verbal | 5 (44) |
| Primarily Non-Verbal | 10 (66) |

To gain a better understanding of the participants enrolled, we asked parents about information regarding their family characteristics. 9/15 families included a stay-at-home parent, and the majority (12/15) of the participants resided in a home with married parents. Only eleven (73%) families reported household income, but among those who did, the distribution of income was similar to the distribution in our city [38].

#### Adherence

Participants spent 149.66 (82.28 – 180.69) minutes robotic walking (Figure 3A) took 7,544 (4,639 – 9,574) steps (Figure 3B) and walked 5 (3.5 – 6) times each week. There were no significant changes in duration (p=0.549), steps (p=0.627), cadence (p=0.135), or number of sessions (p=0.197) across the three months of the intervention. Participants walked for 27.85 minutes (23.9 – 31.8) and 1475 steps (1105 – 1820) per session, with an average cadence of 51.8 (42.3 – 63.9) steps/minute, with the corresponding average weekly usage shown in Figure 3. Just over half (8/15) of the participants met the training requirements and walked at least 150 minutes per/week when averaged across the training period. Of the eight participants who met the training requirements, seven had at least a portion of their intervention period in the spring/summer. Whereas 1/7 who didn’t meet training recommendations had the Trexo in the spring/summer. Further, 6/8 families who met training requirements included a stay-at-home parent who ran the robotic walking sessions. It was believed that participants who were able to train in their home would meet training requirements, however, five of the six participants who trained within their home at least most of the time did not meet training recommendations. It is important to note that only 11% of robotic walking sessions were under 30 minutes, whereas participants trained for less than 5 days a week in 49% of weeks. Three participants did not reach the training recommendations for any weeks during the intervention.

**Figure 3:**
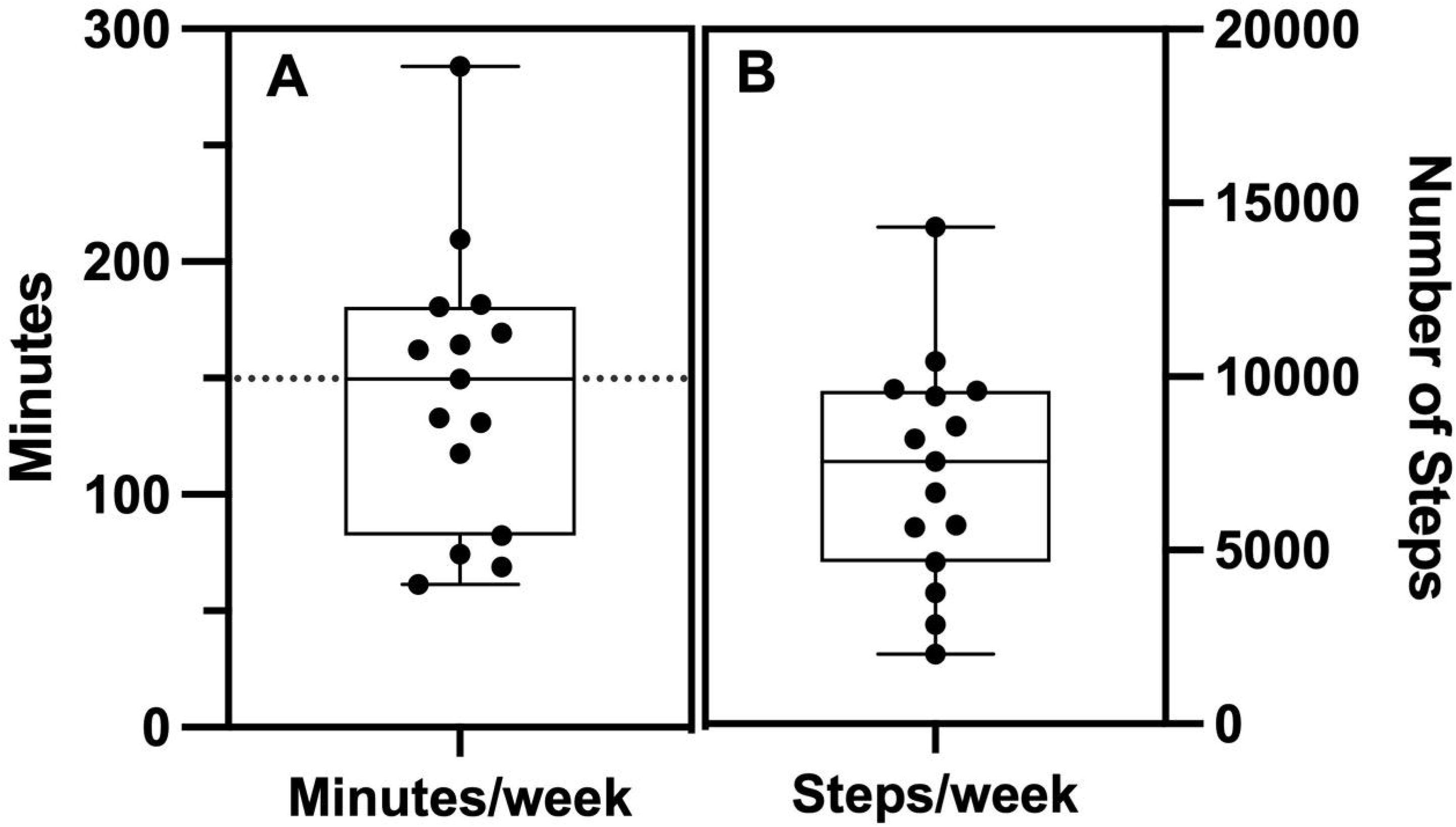
A: Average minutes spent walking each week. Training recommendation was 150min/week; B: Average steps taken each week during training period

Through daily diaries and check-in conversations with parents, we identified barriers to training as prescribed and how many training days this influenced during the intervention period. As discussed above, participants were asked to train at least 5 days/week (corresponding to 71% of days over 12 weeks). Participants did not train on 27 (22-41) days and reported reasons for 19 (8-34) of these days for a total of 281 days non-training with reasons reported by participants. Participants’ reasons for not training, as follows: other activities to attend (20%), didn’t have the time to train (20%), or planned rest days (17%). Other reasons for not training stated by the families were difficulties with the device (8%), weather (5%), no adult available to run the session (5%), illness or surgery (14%), participants not wanting to train (6%) or other reasons training wasn’t feasible that day (5%) (percentages are calculated from the total reported reasons for not training).

#### Acceptability

There were 14/15 parents and caregivers who filled out the daily diaries throughout the training period. Caregivers reported their mood during the robotic walking sessions as well as the child’s mood before and during the sessions on a 5-point Likert scale (Legend: Very Bad = 1, Bad = 2, Okay = 3, Good = 4, Very Good = 5. An increase in mood during robotic walking was reported across all sessions in 9/14 participants. Participants were generally in a “o.k.” or “good” mood prior to training, 3.97 (3.50 – 4.32) and for most did not change or improved by 0.21 (−0.22 – 0.42) on the 5-point scale during training. The lowest rating was 3.52 with most caregivers describing their experience as at least good (4.0 (3.7-4.5)).

#### Practicality

Caregivers spent an additional 13 (10-24) minutes beyond the training time setting up and getting the participant in and out of the Trexo. There was no significant change in setup time throughout the study period (p = 0.097). 88% of the Trexo sessions involved one person running the Trexo session. We did not differentiate whether the remaining 12% required two people or a second person joined for social and/or recreational reasons. Parents most commonly ran sessions (45% of sessions), but aides, school staff, other family, and siblings were also involved in facilitating robotic walking sessions. Sessions were most commonly conducted outside (33%), in school (26%), at home (18%), and in recreation centers (12%).

Families experienced no difficulties in 70% of robotic walking sessions. However, families did express multiple difficulties in 30% of the sessions. Malfunctions with the device occurred in 7% of sessions (see Table 2). The most common malfunctions were the tablet not connecting and the tablet or Trexo battery dying. Additionally, another difficulty was participants not enjoying or uninterested in the robotic walking session (7% of sessions). Other difficulties experienced during robotic walking were troubles with setup and positioning, discomfort during the session, issues using Trexo outside, and limited space. Through the session diaries, it was also tracked why participants did not train.

**Table 2:**
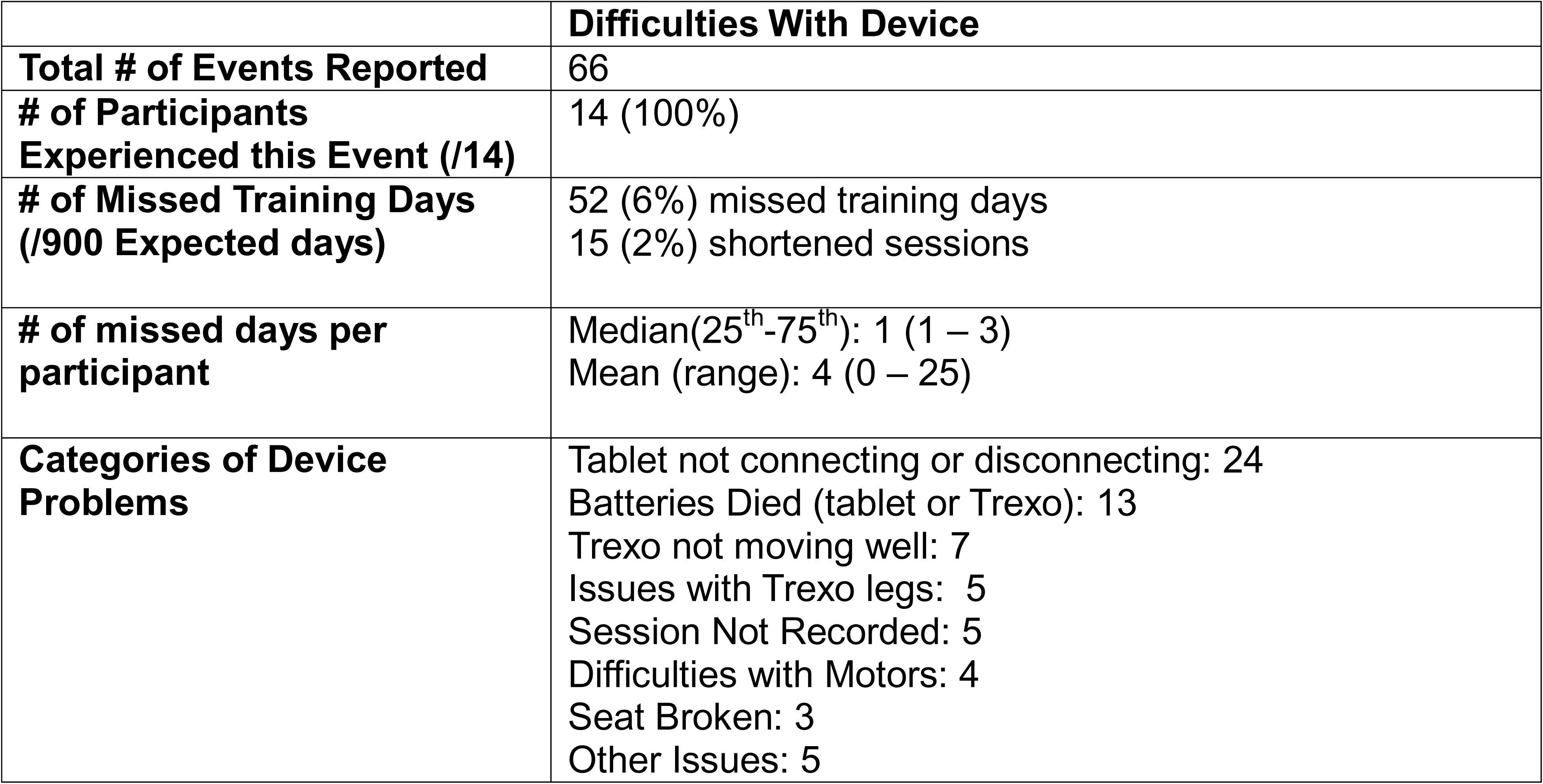
Breakdown of all the difficulties with the device experienced during the intervention period.

|  | **Difficulties With Device** |
| --- | --- |
| **Total # of Events Reported** | 66 |
| **# of Participants Experienced this Event (/14)** | 14 (100%) |
| **# of Missed Training Days (/900 Expected days)** | 52 (6%) missed training days<br>15 (2%) shortened sessions |
| **# of missed days per participant** | Median(25<sup>th</sup>-75<sup>th</sup>): 1 (1 – 3)<br>Mean (range): 4 (0 – 25) |
| **Categories of Device Problems** | Tablet not connecting or disconnecting: 24<br>Batteries Died (tablet or Trexo): 13<br>Trexo not moving well: 7<br>Issues with Trexo legs: 5<br>Session Not Recorded: 5<br>Difficulties with Motors: 4<br>Seat Broken: 3<br>Other Issues: 5 |

#### Adverse Events

Throughout the intervention period, there were 16 minor, and one moderate adverse event related to robotic walking (Table 3). These adverse events impacted 5 participants, and most events were resolved within a day or less. For one participant, a blister resulted in nine missed training sessions, which was the most missed sessions for a single participant. As a result, the participant went to see their orthotist to adjust their ankle-foot-orthoses. The most common event experienced was rubbing on the feet or blisters, followed by bruises on the feet, sore knees, and one participant experienced static electric shock from the motor. Overall, adverse events related to robotic walking led to 11 missed sessions and one shortened session. There were also 30 adverse events that were unrelated to robotic walking; these included injuries and illnesses that resulted in 22 missed training days and affected 11 participants. These adverse events led to 22 missed sessions and 3 shortened sessions.

#### Family Perspective on Feasibility and Logistics

Twelve parents and one participant participated in qualitative interviews with the first author before the intervention started, and nine of those parents and the one participant also did post-training interviews. The participant who took part in the interviews was the adult participant. Many of the children were non-verbal, or the parents felt they would be unable to sit through an interview. Parents were unable to do the second interview for multiple reasons, including scheduling conflicts, being unable to contact parents to book the interview, and their child’s medical needs, making it so the parent no longer had time to participate in the interview. Pre-training interviews took 25 (23 – 29.5) minutes, and post-training interviews took 43 (27 – 46) minutes.

Three themes and five subthemes emerged related to the feasibility and logistics of Trexo use for the parents. The three themes were: Parents enjoyed some Trexo features but found others difficult, logistical concerns surrounding transporting and using the Trexo in the community, and children enjoy Trexo more when compared to traditional therapies. We describe the central meaning of each theme with supporting quotations and interpretations using our theoretical and methodological framework.

##### Theme #1: Parents enjoyed some Trexo features but found others difficult

###### Troubleshooting and technological issues

Some parents described the Trexo as being easy to use and required minimal technological knowledge. Where other parents described that setting up the Trexo was difficult and required an in-depth knowledge of technology. This difference in parent perspectives may be due to parents who had previous technical knowledge finding the Trexo to be quite user-friendly but parents with less technical knowledge found it to be more difficult. Experiencing technical difficulties led to frustration as caregivers had difficulties troubleshooting certain problems and, as a result, had to get in touch with Trexo support. Many parents described having difficulties connecting the tablet to the Trexo with one caregiver stating, “The tablet was having a hard time connecting to the right network, so it’s like having to go in and manually connect the tablet to the Trexo network so the Trexo would talk; that was an issue” [TR03 Dad].

###### Set-Up and Transferring

Some parents found transferring the child in and out of the Trexo easy while others mentioned having difficulties with this. Parents having different perspectives on transferring were typically not impacted by participant size or GMFCS level and varied across participants. Parents mentioned that it would be beneficial if they could get their kid in and out of the Trexo faster with one parent stating, “It would be really nice if there was a way to get him in and out of it a little bit quicker… He walked to school in it every morning… If you’re running 5 min late in the morning, having to get him loaded in and out of that can definitely slow things down but like, I mean, it’s not a big deal,” [TR14 Dad]. Parents specifically discussed how transfers were worse when they were using the Trexo in public places but found them easier when they were at home. However, one mom mention that they didn’t find transfers into the Trexo any more difficult than transfers into the walker, “Transfer in and out of the Trexo was tricky but transfer in and out of the walker is just a tricky thing to do solo in a public place. Because sometimes there’s a stranger that’s like “can I help you?” and I’m like “I don’t think so” but I know I’m not making it look easy” [TR04 Mom].

###### Trexo Features

There were some specific features of the Trexo that caregivers enjoyed; these included: Having the Trexo posterior facing, clipping up the leg buckles was easy, and it’s easy to track usage and steps. There were some other specific features that families mentioned they did not enjoy: The Battery was heavy, the wheels got stuck on different surfaces (i.e. outside, carpet, etc.), the height of the Rifton could not be adjusted with just one person, and clipping up the footplates was difficult. When discussing the design of the Trexo, one mom stated, “I don’t want to be so harsh as to say like poorly designed, but there could be some pretty significant tweaks that would make it easier on parents. Like my fingers are still so chewed up from like the buckles and reaching around like yeah that was difficult” [TR11Mom]. Some parents noticed that some days the Trexo looked more awkward, and the steps did not look as fluid, leading families to feel like the Trexo did not work the same day to day.

##### Theme #2: Logistical concerns surrounding transporting and using the Trexo in the community

###### Where families used the Trexo

All parents mentioned some difficulties with logistics when trying to use the Trexo at home and in different areas of their community. Parents stated that they loved the opportunity for their child to use the Trexo at school, as this took some of the stress off them when it came to running the sessions. One mother stated, “He wouldn’t walk in school before the Trexo… He quite enjoyed the time, so the benefit of sending it to school was that he did get the experience of walking around in school and showing his friends and all of that. So that was the one real benefit.” [TR03Mom]. Some parents loved the ability to use the Trexo at home, where it just wasn’t feasible for others due to their house being too small. As a result, many families used the Trexo outside instead. However, weather and other difficulties with the Trexo did get in the way of using it outside frequently. When talking about the difficulties with using the Trexo outside, one dad stated, “… the wheels are terrible. When we did go outside in the winter, when it was kind of clear enough, they would get all clogged up with rocks and whatever else, and I mean it really was another major limiting factor.” [TR18 Dad].

###### Transportation

Parents found the Trexo very difficult to transport and found this was impossible to do with only one caregiver, thus requiring them to organize additional help whenever they wanted to use the Trexo in a public setting. Parents felt that the Trexo is too “big and bulky” [TR11 Mom]. One parent stated that their biggest difficulty was “Just where we could do it… I could run it by myself, but if I needed to take it somewhere, I still had to get another person to come help me ‘cause… I can’t lift it; it’s too heavy” [TR01Mom]. This led to caregivers feeling stressed out due to there being so many logistics to figure out when trying to use the robotic walker in different areas of their community.

##### Theme #3: Children enjoy Trexo more when compared to traditional therapies

The majority of parents discussed how their child enjoys the Trexo more than typical rehabilitation. This relieved parental stress because they no longer had to go to appointments or force the child to do something they did not enjoy. All parents discussed how their child does not enjoy using their standing frame, and the Trexo was a great tool to replace that, and the children actually enjoyed it. One dad discussed how he used to worry that he was doing something wrong when he helped facilitate rehabilitation at home, but with the Trexo, “The machine takes care of itself and makes sure that it’s doing what it’s supposed to do.” [TR14Dad]. One mom did discuss how “[TR11 name] responds better to other people I don’t think he would have put up as much of like all of the arguments in fighting if it was somebody else someone getting him into the Trexo… it definitely makes it more difficult, and I think stressed our relationship at certain times right” [TR11Mom]. However, for the majority of children in this study, they enjoyed the Trexo and were more engaged than in other forms of therapy.

### Family Goals and Impacts

#### Canadian Occupational Performance Measure (COPM)

Parent perceptions of performance (p = < 0.001) and satisfaction (p = < 0.001) increased significantly during the Trexo training period (Figure 4). Before training, performance scores were 3.4 (2.6 – 5) and increased to 7.0 (5.4 – 7.8) following the intervention period. Satisfaction scores were 3.4 (2.6 – 3.9) before training and 7.8 (5.9 – 8.8) following the intervention period. In the follow-up period, performance scores decreased in 7/13 of the participants and satisfaction scores decreased in 8/13 participants, though decreases were not significant. All participants had goals related to functional mobility. The other COPM goal categories were as follows: personal care (11/15), socialization (7/15), active recreation (4/15), school &/or play (4/15), quiet recreation (3/15) (Figure 5).

**Figure 4:**
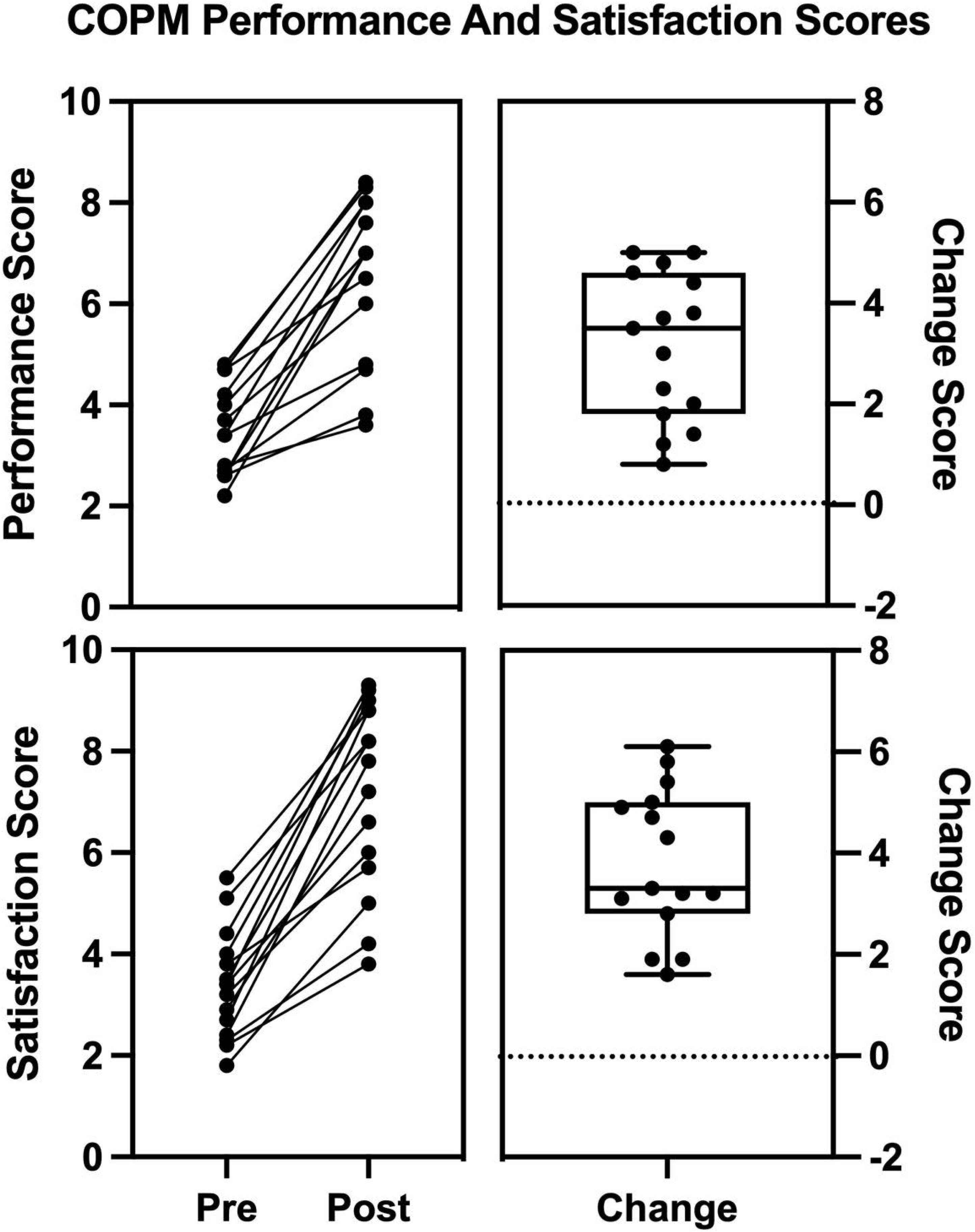
COPM performance and satisfaction scores before and after the intervention period and change in scores between these two timepoints. There was a significant change in both performance (p=<0.001) and satisfaction (p=<0.001).

**Figure 5:**
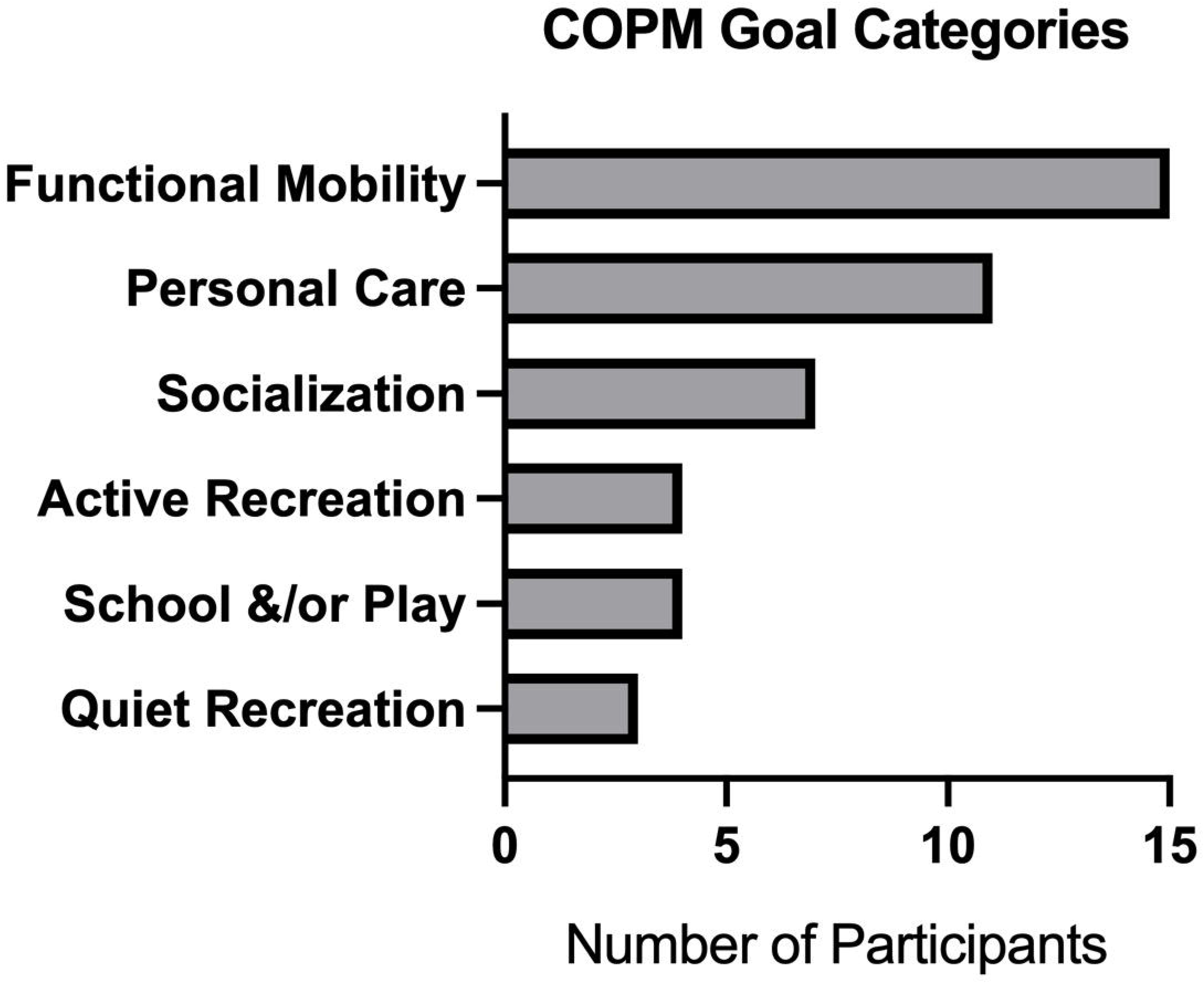
COPM goals were classified into categories, to understand which categories parents most commonly wanted to see improvement in. With functional mobility being the most common category.

#### Quality of Life Questionnaires

There was no significant change in any of the quality-of-life questionnaires. Both the EQ-5D-y health dimension score (p = 0.158) and health ratings (p = 0.195) did not improve significantly. Further, there was no change seen in the Carer-QoL Questionnaire (p = 0.856). There was no significant change in CP-CHILD before and after the intervention. The most improvements were seen in the personal care and activities of daily living domains, though not significant.

#### Family Impacts and Perspectives

When discussing the impacts robotic walking had on families, three main themes and five subthemes were identified: Families bonded and were able to interact in a new way, parents’ perspectives on their child changed, and parents noticed that they had more positive social interactions while their child was walking in the Trexo.

##### Theme #1: Using the Trexo changed the parents’ perspectives on their child

###### New Expectations for their child’s future

Using a robotic walker in their home community was a new experience for all the families in this study. This experience changed how parents view their child, and they realized there were more possibilities for their child’s future than they may have initially thought. Seeing their child use the robotic walker made parents feel like their child’s diagnosis was less scary, because this experience opened their eyes to the possibilities of technology and how it may positively impact their child’s future. “I have no doubt in my mind that he’s gonna be walking around by himself one day… with the assistance of something. [He’s] not gonna be bound to a wheelchair… So one thing that I really liked about this is being reassured that the future is coming for him. And with where technology is, it’s gonna be a lot better than somebody in his situation 10 years ago” [TR14 Dad]. Furthermore, parents also mentioned that due to robotic walking, their child was now exceeding the expectations that doctors originally set for their child. Seeing the participant in the Trexo also changed other family members’ views on what the participant is capable of. One mom was discussing when the participant’s grandma saw her in the Trexo and mentioned, “Yeah, my mom is like, ‘wow the sky is the limit for this girl’, she’s got a lot of potential that hasn’t been taken care of” [TR04 mom].

###### Changes in Independence

A few parents believed the Trexo gave their child more independence, whereas other parents whose child typically uses a power wheelchair for mobility felt like the Trexo gave their child less independence. One mom mentioned, “Also like the lack of like independence like he can’t do any of it yet on his own he could be right there” [TR11 Mom]. As a result of the participants exceeding doctors’ expectations and gaining independence, parents’ views changed, and they realized their child may be more capable than they may have originally thought. However, it is important to note that two parents mentioned seeing no change in their child.

##### Theme #2: Families bonded and were able to interact in a new way

###### Changes in participants’ mood

This experience positively impacted the participants’ moods and changed how they interacted with their family members. When discussing robotic walking one mom mentioned, “As a parent being able to see her joy that she can actually say I’m going to walk to the stop sign and back is unbelievable she never been able to do that so she’s gone from really a girl in a wheelchair to a walking girl. So it’s like something that we hoped for and longed for, and now she’s actually doing it” [TR01Mom]. One dad mentioned that he experienced a decrease in parental stress due to “His attitude as well. Right, you know again. There are times where you know he could be sour… just have a not good day, and those have seemed to be fewer and far between with us.” [TR14 Dad]. This led to parents and their child having more positive interactions. One mom did mention that their child did not enjoy using the Trexo, and that caused parental stress and put a strain on their relationship.

###### Walking allowed for bonding and changed conversations between the parent and child

Having the families lead the robotic walking sessions created a new way for family members to bond with the participant. All parents mentioned being able to walk beside and hold hands with their child for the first time in their lives. It was the first time in their child’s life that parents were able to have this experience with their child, and it excited the parents to be able to experience something that most parents can experience daily. Parents mentioned how their child opened up more on Trexo walks. When describing her walks with her son, one mom mentioned, “We had really good conversations on our walks… Without a lot of prompting, he would tell me about his day so our conversations when he was walking in the Trexo… he starts to open up and tell me about his day which he doesn’t like doing like right after school or whatever, so he was doing that in the Trexo.

###### Sibling Bonding

Siblings were able to help with the Trexo and walk with their sibling, which was a new way they were able to interact with their sibling. Siblings also liked to entertain the participants to keep them enjoying the robotic walking session. One mom mentioned how “[TR04 sister] can independently run her around in the Trexo as a [school aged child] which is pretty cool, like we help her get in and then she’s like “I’m gonna take my sister for a walk you don’t come” and take her around so that’s kind of cool that she’s [school aged] and it’s nice to have time that’s not with adults, even though adults are nearby and accessible and it’s safe” [TR04 Mom]. Having the opportunity to see their children have fun together and interact was really important to the parents. While robotic walking did bond a lot of the families, it is important to mention that one parent stated that their child did not like going in the Trexo, which put a strain on the parent-child relationship. It was also difficult for some families to get their whole family out for a walk, so not all family members participated in the robotic walking sessions.

##### Theme #3: Parents noticed that they had more positive social interactions while their child was walking in the Trexo

One of the biggest impacts parents noticed was a positive shift in social interactions and changes in how the parents and participants interacted with people in their home community. Parents noticed that peers were treating the participant differently when they were in their Trexo. One mom mentioned how she often feels like peers ‘facilitated her child’s disability’ but when he is in the Trexo they treat him like he has a fun new toy, and he was super excited to be able to show off the Trexo. This idea of showing off the Trexo was mentioned multiple times between families. The participants became excited and proud when their peers were able to see them robotic walking. Participants were much more comfortable getting attention while robotic walking versus the attention they typically get in their wheelchairs. While in public with a child with a disability, parents often felt like people, “…just stare at us or like just ignore us, I don’t really think it makes a difference but if they just stare at us that it’s uncomfortable. If you have a question, just come ask… I think that was the one neat thing with the Trexo is that people were more willing to come up and talk to us instead of just stare” [TR03 Mom]. Many families enjoyed this and welcomed the new discourse around their child’s disability with the public. Though the public interactions did feel a little overwhelming for some families, “All like that are just asking way too many questions… in front of [TR14 name] that’s a lot of stuff that I don’t talk with him about, necessarily right, [and the] the Trexo puts a big target on you for attracting people like that.” [TR14Dad].

## Discussion

This study provides promising evidence showing that robotic walking in the participants’ home communities resulted in benefits relevant to families (ie, their reason for doing the intervention). Families noted logistical challenges, and only half reached the training recommendation (150 minutes/week). Nevertheless, all reported improvements in performance and satisfaction with goals, and many mentioned additional benefits to the family and participant.

Though we tried to recruit a wide range of participants representative of those who may use robotic walking aids, there were some patterns that may impact generalizability. The majority (12/15) of our participants functioned at GMFCS level IV. Only a few participants (3/15) lived with a single parent, and most had a stay-at-home parent (9/15), which may have made this study more feasible, as one parent was more available to help with the Trexo. It is worth noting that it’s more common for children with a disability to have a stay-at-home parent[39] Our study does include participants with diagnoses beyond cerebral palsy and a wide range of ages, which is representative of children currently using the Trexo [20]

The frequency of training (5 days/week) requested in this study was clearly difficult and not achieved in almost half of all weeks of training, whereas the duration (30 minutes/week) was achieved more consistently (89% of training sessions). There is no known optimal training dose or even consensus for robotic walking to result in a clinical impact [40]. Other studies involving the same device have very different training amounts, with those being 240 −300min/week (1 hour a day, 4-5 days/week) or 60 mins per week (30min/day, 2 days/week) [41], [42]. The rehabilitation principles that guided this study suggest that frequent training for at least 30 minutes is needed to stimulate neuroplastic changes [22]. While we did not measure neuroplasticity directly, even participants who trained only 2.5x/week saw improvements in performance and satisfaction with rehabilitative goals. This is consistent with a study involving low doses of robotic walking (30-45 minutes twice a week) that found improvement in walking [43].

Multiple logistical challenges contributed to the difficulty in meeting training recommendations. Transportation was one of the major barriers mentioned when families were trying to use the Trexo around their home communities. Many individuals living with a disability experience challenges with transportation in their daily lives, making it difficult for them to access many opportunities [44]. Therefore, we would recommend that families and clinicians who are interested in using the Trexo discuss transportation, and companies consider portability when designing robotic walking devices. Being able to use the Trexo outdoors clearly facilitated training more frequently, though the weather and our winter climate were limiting factors. The time of year had an impact on how frequently families were able to robotic walk. These logistical challenges had a much larger impact on training than adverse events. It’s important to note that adverse events related to the intervention were experienced in <1.5% of sessions. The most common adverse events experienced were abrasions and rubbing on knees and feet, which are common adverse events experienced in other activity-based interventions [45]. While inclusion criteria were set to ensure participants were safe while robotic walking (eg, no weight-bearing or physical activity limitations), our population (individuals GMFCS III-V) have a higher prevalence of conditions that may increase the risk of adverse events[45]. Future studies could consider a gradual onset of training to build up endurance for weightbearing on the feet. Participants may also need to obtain new ankle-foot-orthoses & shoes with the expectations of increased stepping and weightbearing. Though there were limitations experienced, all children experienced benefits related to robotic walking.

Everyone saw improvements in the performance and satisfaction with goals. This universal experience of perceived benefits highlights the importance of using personalized goals to ensure the measurements used are relevant to outcomes important to families [46]. Yet, this is infrequently done in the robotic walking field, as is highlighted by recent systematic reviews [13], [18], [47], [48]. While goals were diverse, all participants had at least one goal related to functional mobility, which is a domain with documented benefits [18]. There were also additional benefits observed in performance and satisfaction of goals in other areas where parents hoped to see improvements, and other benefits families noted that were not necessarily anticipated in advance.

Most parents discussed robotic walking having a positive impact on their family, despite the potential for added stress from having families run the intervention. However, there were no significant changes in measures of quality of life used in this study. Finding a reliable quality of life questionnaire for children with cerebral palsy can be difficult, and using parent proxies may not accurately represent the quality of life of the child [49]. In our study, parents anecdotally mentioned that the tools we used do not accurately represent the lived experience of their child. Though there was no significant change in quality of life, parents mentioned that they noticed an increase in family bonding as a result of the robotic walking intervention. Having parents and siblings run the robotic walking sessions led to parents and siblings bonding with the participant in a new way. It can be difficult for families with a child with a mobility impairment to find time and an avenue to do activities together, but for these families, robotic walking has provided this. Family bonding and support have been associated with better psychological and social outcomes [50]. It has also been noted that siblings enjoy the opportunity to support their sibling in physical activity endeavours [51], [52]. Further, this experience of family bonding would not occur while using a robotic walker in a clinical setting. Many children do not enjoy typical rehabilitation activities done at home, such as using a standing frame and stretching, but many children enjoyed using the Trexo. This led to more positive interaction between parent and child and decreased parental stress. The benefits seen in this study extend beyond the family unit, as many parents mentioned a positive change in social interactions.

One main impact experienced was a positive change in social interactions while in community settings. Parents with children with a disability often feel like they receive stares and judgmental looks while in public with their child [53]. Poor social interactions can impact the self-esteem and mental health of children living with a mobility impairment [54]. While robotic walking in community settings, parents mentioned that they now get fewer judgmental looks and people are more interested in interacting with their child. These observations are similar to walking with a manual walker in community settings by those who are able [9]. As none of our participants use a walker in community settings, they would not be able to experience these benefits without the use of the robotic walker.

## Conclusion

The results of this study demonstrate that family-led robotic walking may be feasible, though we need a better understanding of the minimal training volume required to see benefits. Even though participants trained as little as half of our recommended 30 minutes/day, 5 days/week, both performance and satisfaction in preset personalized goals increased for all participants. Difficulties with the device and other logistical concerns were mentioned as a major reason for not reaching training recommendations. This single-group study design was appropriate for an initial investigation of this device and will enable to exploration of more impacts in a larger number of participants and more controlled studies. This study represents the largest and most diverse investigation to date of an untethered robotic walker in a mixed population of children, youth, and young adults. It provides groundbreaking work for future studies regarding the impact on the health and well-being of individuals with childhood-acquired disability.

## Acknowledgements

We would like to thank the participants and patient partners who guided this project: Sarah Best, Kathleen Best, Dario Facca, Maggie Facca, and Erin Hannigan. Funding for this trial was provided by the Alberta Children’s Hospital Foundation’s Vi Riddell Centre for Pediatric Pain and Rehabilitation. This Project was also supported by Kids Brain Health Network, with the financial support of Health Canada, through the Canada Brain Research Fund, an innovative partnership between the government of Canada (through Health Canada) and Brain Canada. Additionally, the first author would like to thank NSERC BRAIN CREATE for providing salary support.

## Declaration of Interests

Trexo Robotics provided in-kind support, including loan of devices, training and support, as well as access to their data on the device usage. A data-sharing agreement ensures the academic independence of all of our work done with the loaned devices. They have not participated in this manuscript’s preparation. Our research team has also received an unrestricted donation from Trexo Robotics ($40000 in 2023). The authors have no further conflicts of interest to disclose.

## Data Availability Statement

Access to the full dataset supporting the findings of this publication is available upon reasonable request from the following repository https://doi.org/10.5683/SP3/538NWV. A publicly accessible subset of the data can be found at https://doi.org/10.5683/SP3/B5FZZA. Due to the presence of information that could compromise participant privacy, the complete dataset is not openly available.

## Notes

### Clinical Trial

NCT05473676

### Funding Statement

Funding for this trial was provided by the Alberta Children's Hospital Foundation. This project has also been made possible by Kids Brain Health Network, with the financial support of Health Canada, through the Canada Brain Research Fund, an innovative partnership between the government of Canada (through Health Canada) and Brain Canada.

### Author Declarations

Conjoint Health and Research Ethics Board at the University of Calgary (REB21-1166)

